# LSD600: the first corpus of biomedical abstracts annotated with lifestyle–disease relations

**DOI:** 10.1101/2024.08.30.24312862

**Authors:** Esmaeil Nourani, Evangelia-Mantelena Makri, Xiqing Mao, Sampo Pyysalo, Søren Brunak, Katerina Nastou, Lars Juhl Jensen

## Abstract

Lifestyle factors (LSFs) are increasingly recognized as instrumental in both the development and control of diseases. Despite their importance, there is a lack of methods to extract relations between LSFs and diseases from the literature, a step necessary to consolidate the currently available knowledge into a structured form. As simple co-occurrence-based relation extraction (RE) approaches are unable to distinguish between the different types of LSF-disease relations, context-aware transformer-based models are required to extract and classify these relations into specific relation types. No comprehensive LSF–disease RE system existed, primarily due to the lack of a suitable corpus for developing it. We present LSD600, the first corpus specifically designed for LSF-disease RE, comprising 600 abstracts with 1900 relations of eight distinct types between 5,027 diseases and 6,930 LSF entities. We evaluated LSD600’s quality by training a RoBERTa model on the corpus, achieving an F-score of 68.5% for the multi-label RE task on the held-out test set. We further validated LSD600 by using the trained model on the two Nutrition-Disease and FoodDisease datasets, where it achieved F-scores of 70.7% and 80.7%, respectively. Building on these performance results, LSD600 and the RE system trained on it can be valuable resources to fill the existing gap in this area and pave the way for downstream applications.

## Introduction

Diseases are influenced by both lifestyle and genetic factors (1–4). Substantial evidence suggests that targeted lifestyle interventions can significantly enhance disease prevention and management in precision medicine (5,6). Considering the growing importance of lifestyle factors (LSFs) in disease, attention has been placed towards mining the literature for such relations (7–9). Despite these efforts to extract and centralize scientific knowledge, the focus is almost exclusively on the relation between diseases and nutrition, and the relations between other LSFs and diseases remain largely unconsolidated and scattered across the scientific literature. To mine LSF–disease relations from the scientific literature, the first step is to detect mentions of these entities in text. Then a straightforward approach to identify pairs of related diseases and LSFs is co-occurrence-based Relation Extraction (RE). While this will allow the detection of related entities, the nature of their relation will remain unknown. Although this is a non-issue for gene-disease relations (10), as their nature is mostly causal, the case is completely different when it comes to LSF–disease relations. For example, ‘exposure to radiation’ is well known to be harmful to health (11), causing diseases like skin cancer (12). At the same time, in modern medicine, radiotherapy, which also exposes the human body to radiation, is a common method for treating cancer (13). Therefore, simple RE approaches cannot capture the complexity of LSF–disease relations and it becomes essential to develop more sophisticated approaches that can distinguish between different types of relations — from simple statistical associations (either positive and negative), to more concrete causal, or preventive relations.

The advent of transformer-based language models such as BERT (14) and RoBERTa (15) has revolutionized the field of RE, including domain-specific areas like biomedical RE (16). However, to optimally use these transformer-based models, dedicated corpora for fine-tuning are required. The focus of the BioNLP community regarding RE corpora for disease entities has been primarily on disease–gene (17,18) or disease–chemical (BC5CDR) relations (19). When it comes to LSF–disease relations, only two relevant corpora exist, namely the ND (Nutrition and Disease) corpus (7) and the FoodDisease corpus (9). However, there is a notable lack of comprehensive corpora that cover a broader range of lifestyle factors beyond nutrition and different types of LSF–disease relations, highlighting the need for dedicated datasets for training LSF–disease relation extraction models.

In this study, we introduce LSD600, the first corpus specifically focused on LSF–disease relations. LSD600 consists of 600 abstracts annotated with LSF–disease relations, encompassing 1900 relations covering eight different relation types and nine categories of lifestyle factors (20). We have used LSD600 to train a transformer-based model on the multi-label LSF–disease RE task, which achieved an F-score of 68.5% on our held-out test set. We further validated the model on two independent external corpora of nutrition–disease relations and saw comparable performance. This model represents a first important step in extracting LSF–disease relations from biomedical literature, which can form the basis for knowledge graphs.

## Materials and Methods

### LSF–disease relation corpus

#### Document selection for annotation

Given that most of the over 37 million scientific articles available in PubMed do not pertain to LSF–disease relations, this project requires a rigorous selection process to identify relevant abstracts where such relations are expected to appear.

We started with the 200 abstracts from the LSF200 corpus (20), which were pre-annotated with a wide spectrum of LSF named entities. However, since this corpus was compiled to develop NER systems for LSF detection, many did not contain disease mentions or LSF–disease relations.

We therefore selected 400 additional abstracts from PubMed, which were required to mention at least five LSF and five disease mentions, based on using the JensenLab Tagger (https://doi.org/10.1101/067132) and publicly available LSF (20) and disease (10) dictionaries. Considering the uneven distribution of LSFs in the scientific literature (20), with terms related to nutrition being more common than terms from other categories, we selected at least 30 documents for each LSF category. The final corpus consists of 600 abstracts and is named LSD600.

#### The Relation Types Schema

To capture the spectrum of relations between LSFs and diseases we defined eight different relation types:

1. ***Statistical Association***: Statistically significant association between two entity types, requiring the existence of an appropriate control group and the implementation of a statistical test.
2. ***Positive Statistical Association***: Subclass of ***Statistical Association*** where a positive effect is clearly stated.
3. ***Causes***: Subclass of ***Positive Statistical Association*** where causality is clearly implied.
4. ***Negative Statistical Association***: Subclass of ***Statistical Association*** where a negative effect is clearly stated.
5. ***Controls***: Subclass of ***Negative Statistical Association*** where a beneficial impact of the LSF on the disease is stated.
6. ***Prevents***: Subclass of ***Controls*** where the LSF hinders the disease from occurring.
7. ***Treats***: Subclass of ***Controls*** where the LSF has a therapeutic effect on the disease.
8. ***No Statistical Association***: Relations where the absence of statistical association is clearly stated in text.

All relation annotations are non-directional, since in associative relations the direction is undefined, while in relations of impact it is self-evident. For more details and examples on each relation type, please refer to our annotation documentation on Zenodo and Figure 1.

**Figure 1.**
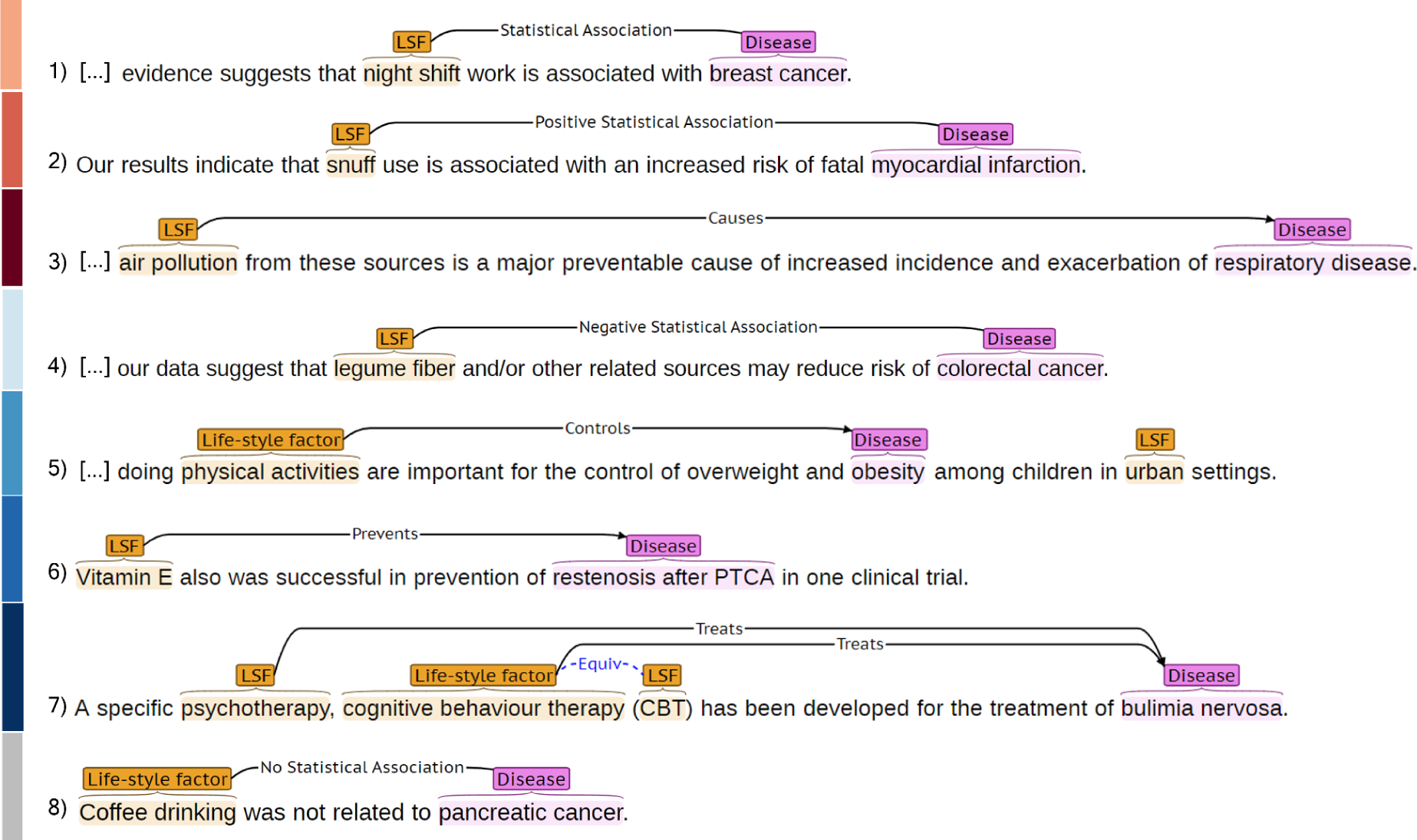
Illustration of the eight LSF–disease relation types in LSD600. Each line and associated color corresponds to a different type of relation: (1) *Statistical Association*, (2) *Positive Statistical Association*, (3) *Causes*, (4) *Negative Statistical Association*, (5) *Controls*, (6) *Prevents*, (7) *Treats*, and (8) *No Statistical Association*. For each relation type, we show an example as it looks in the BRAT RapidAnnotation Tool.

#### Named entity and relation annotation

We pre-annotated LSD600 with disease and LSF entities using the dictionary-based JensenLab tagger (https://www.biorxiv.org/content/10.1101/067132v1). As LSF200 is a corpus with LSF annotations, we only used the tagger to add disease pre-annotations to those abstracts. All pre-annotations were subsequently manually checked and corrected, and relations between them were manually annotated on all 600 abstracts. We systematically annotated equivalent entities, which is essential for evaluation purposes (21). Figure 1 illustrates entity and relation annotation using an example for each relation type.

#### Manual annotation process and corpus evaluation

High-quality, consistent annotations are a prerequisite for good performance when fine-tuning deep learning classifiers, hence a clear set of guidelines is crucial (22). Our annotation guidelines were formed and updated during a first round of Inter-Annotator Agreement (IAA) after three researchers individually annotated 30 abstracts and discussed their inconsistencies. A second round of IAA was performed with a fourth annotator, who is an expert on the field of lifestyle factors, to further update and solidify the final guidelines. To evaluate the quality of the guidelines and the corpus, an F1-score was calculated.

We annotated relations across sentences and made sure to include both direct and indirect relations between entity types. For a statistical association to be annotated, it must be statistically significant and the existence of an appropriate control group must be stated. To determine if an association is positive or negative, we considered both textual descriptions and any stated odds ratios, risk ratios, hazard ratios, or coefficient values. Hypothetical statements are not annotated as either.

The BRAT Rapid Annotation Tool (23) was used to perform all the annotations.

### Transformer-based relation extraction

#### Relation extraction pipeline

The transformer-based system employed for relation extraction was adapted from a previously developed binary relation extraction tool, which has proven effective for RE in past applications (24,25). As described in these earlier papers, we frame RE as a multi-label classification task, aiming to determine which relation types (if any) exist for a pair of candidate named entities in the text. The system uses an architecture comprising a pre-trained transformer encoder and a decision layer with a sigmoid activation function. It can leverage pre-trained language models and accepts training, validation, and prediction data in both BRAT standoff and a custom JSON format, supporting extensive hyper-parameter optimization. Evaluation metrics are computed after each training epoch for hyper-parameter tuning. The system is trained for a predefined number of epochs, choosing the model weights that gave the highest F1-score.

As no separate sentence boundary detection is used, the system can train on and predict cross-sentence relations at the document level. To inform the model about which pair of named entities in the text to predict relation labels for, the entities are replaced by “unused” tokens from the model. This masking approach prevents the model from learning based on the actual entities rather than the surrounding context. For additional details on data preprocessing and the pipeline, please refer to (24)

#### Experimental setup

In our experiments, we partitioned the LSD600 corpus into distinct training, development, and test sets. The system was trained on the training set, and hyperparameters were optimized by grid search to obtain the best F1-score on the development set. The test set was used only once for prediction and evaluation of the best model, after determining the optimal hyper-parameters.

#### Validation on external corpora

In addition to evaluating our model on the held-out test set from LSD600, we validated it on two external corpora, namely the ND (Nutrition and Disease) corpus (7) and the FoodDisease corpus (9).

An overview of these corpora is provided in Table 1.

**Table 1.** Overview of external corpora used for RE system validation.

| Corpus | Nutrients | Diseases | Relations | Sentences | Relation Types |
| --- | --- | --- | --- | --- | --- |
| ND | 234 | 278 | 513 | 1218 | *Related, Unrelated* |
| FoodDisease | 608 | 608 | 464 | 608 | *Cause, Treat* |

The relation types in these corpora are not completely aligned with LSD600. To calculate performance of our model on these corpora, we mapped the relation types of the model to their relation types. The ND corpus uses only *Related* versus *Unrelated* labels. We collapsed all subclasses of *Statistical Association* in our hierarchy into one category, which can be mapped to *Related*. *No Statistical Association* along with no extracted relation is considered as *Unrelated*. For the FoodDisease corpus, the existing relation types are called *Cause* and *Treat*; however, these do not correspond to our classes with the same name. Instead, we collapsed all subclasses of *Positive Statistical Association* and mapped it to *Cause*. Similarly, we collapsed all subclasses of *Negative Statistical Association* and mapped it to *Treat*. Since we do not retrain the model, we evaluate it on the entirety of both corpora.

## Results and discussion

### Corpus statistics

As its name suggests, LSD600 comprises 600 abstracts annotated with LSF–disease relations. Unlike most existing RE corpora, we do not limit the annotations to intra-sentence relations, with 16% of relations in LSD600 being cross-sentence. This should be compared to the less than 5% observed in several other biomedical RE corpora (24–28). Despite the inherent difficulty of cross-sentence relation annotation, the final IAA F1-score was 82.1%, demonstrating that the annotation quality of LSD600 is high.

The 600 abstracts that make up the corpus were randomly partitioned on the document-level into a training set (60%), a development set (20%), and a held-out test set (20%). The corpus contains a total of 1900 manually annotated relations, which are distributed over eight relation types that are organized into a hierarchy. The breakdown of LSF–disease relations across corpus partitions and relation types is shown in Figure 2. The by far most frequent relation type was *Positive Statistical Association* (32%), which reflects that the literature primarily focuses on lifestyle risk factors and that it is harder to demonstrate a causal relation.

**Figure 2.**
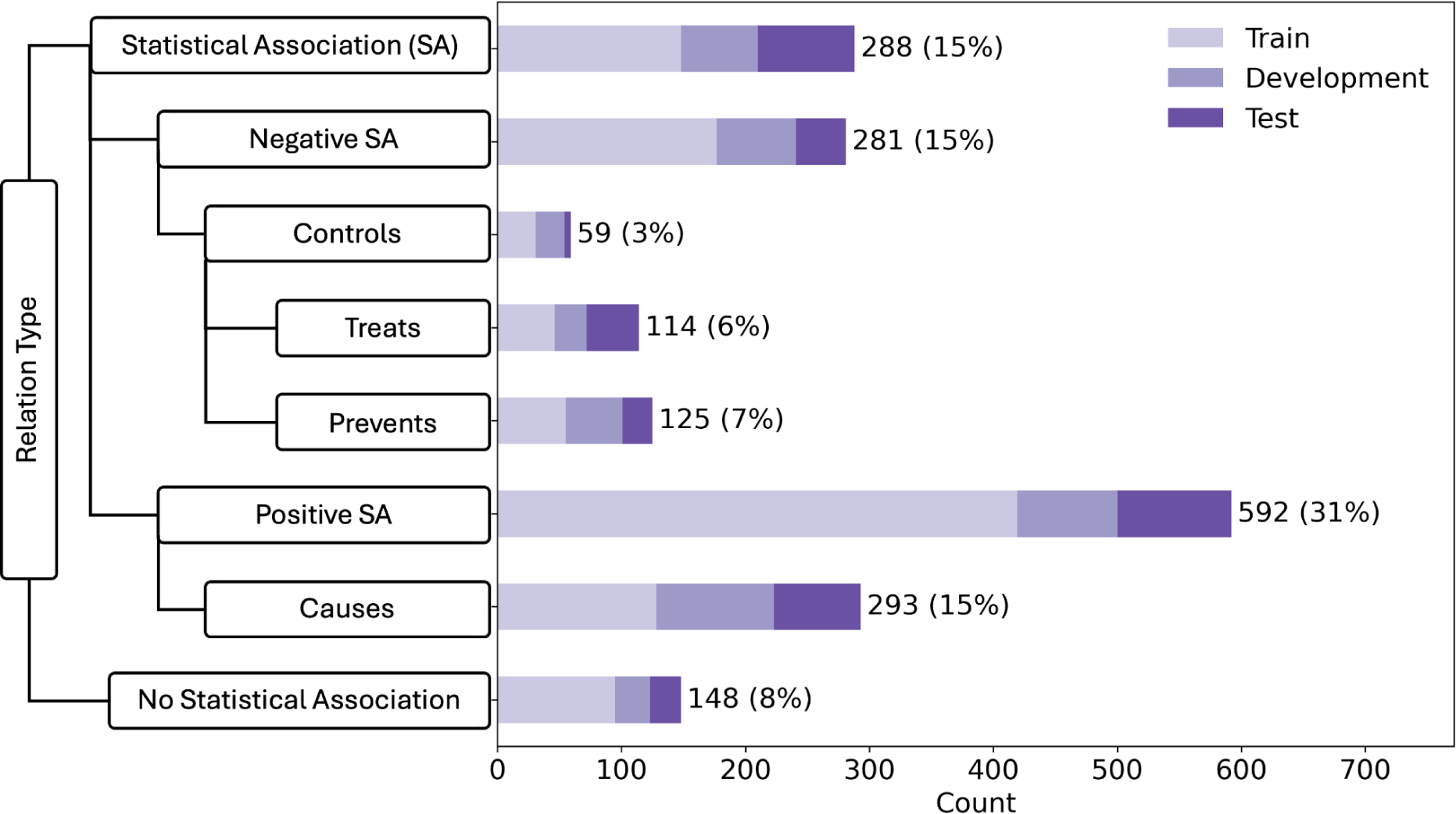
LSD600 statistics for the eight distinct LSF–disease relation types. The left side of the figure shows our relation schema, emphasizing superclasses and subclasses. The bar chart on the right side shows the number of examples for each relation type in the corpus. We distributed the corpus into train:development:test sets in a 60:20:20 ratio, with each bar showing the breakdown for each relation in the different sets.

The 1900 relations in LSD600 involve 5,027 diseases and 6,930 LSF entities. Figure 3 provides an overview of the distribution of relations per abstract, while Figure 4 shows the distributions of disease and LSF mentions per abstract. Additional details are available in Supplementary Tables 2,3 and 4.

**Figure 3:**
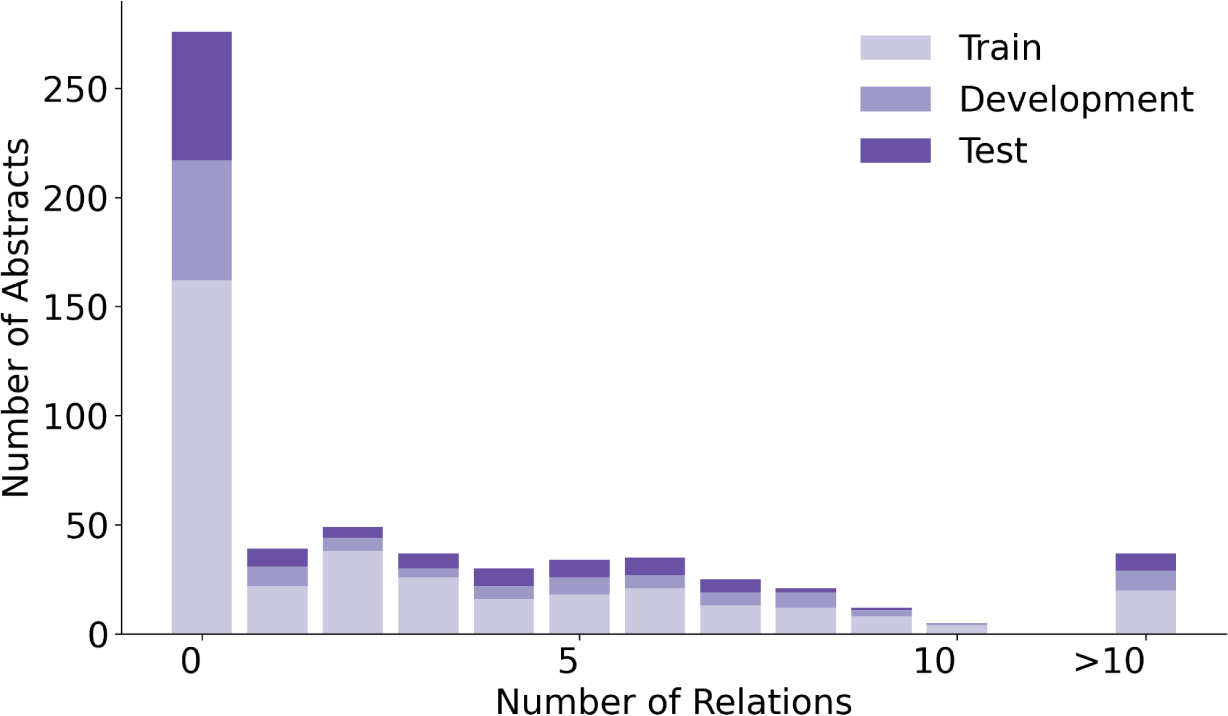
An overview of the distribution of relations in LSD600. The bar chart shows how many abstracts contain how many annotated relations in the training, development, and test sets.

**Figure 4:**
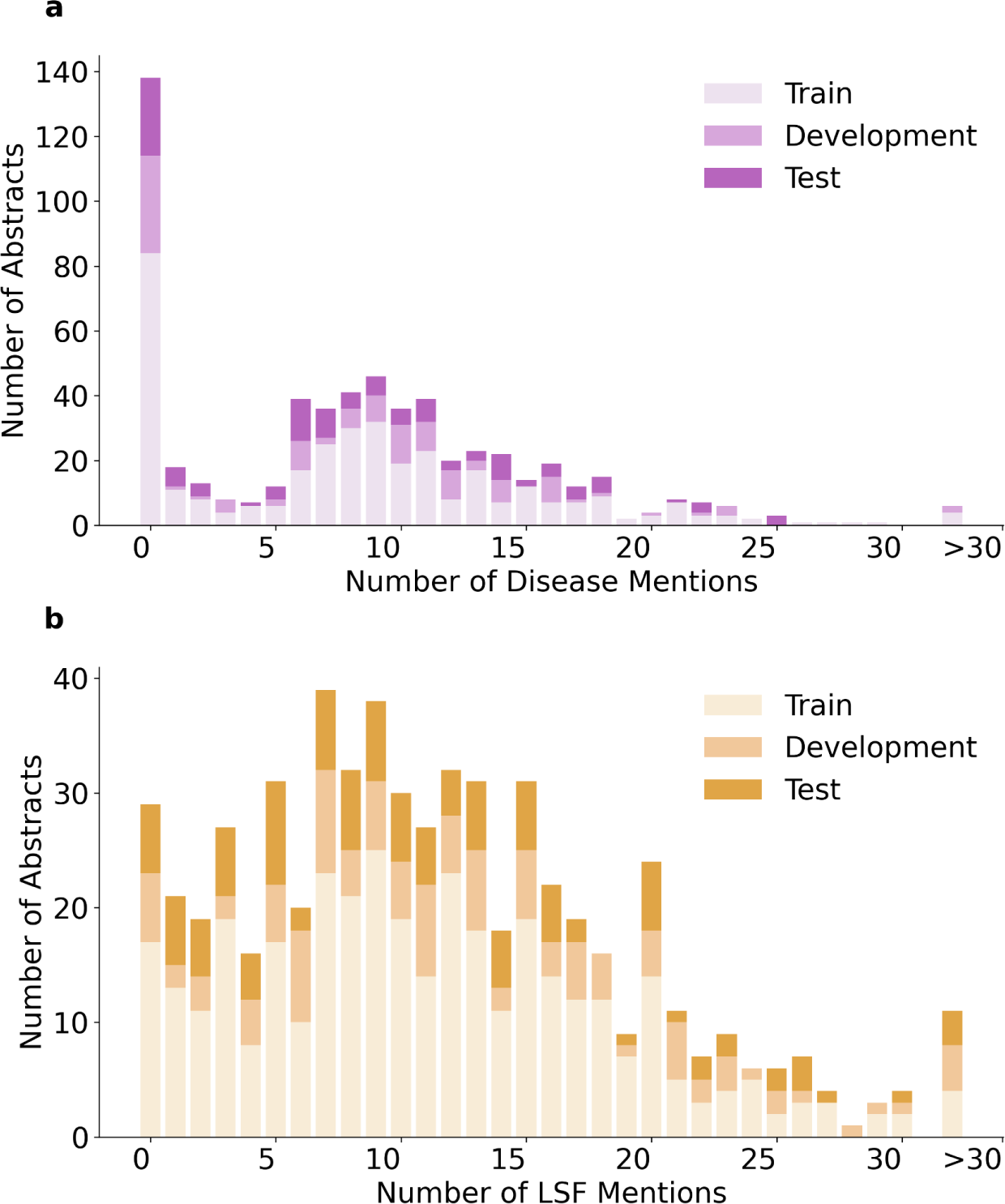
Distribution of Disease and LSF mentions in LSD600 abstracts. (a) Bar chart showing how many abstracts contain how many disease mentions. (b) Bar chart showing how many abstracts contain how many LSF mentions. Bars are divided into train, development, and test sets.

In 276 abstracts, no relations were annotated. This outcome can be primarily attributed to two factors: First, our guidelines specify that we only annotate relations that provide solid, factual information. Second, there are 138 abstracts with no annotated disease mentions. 137 out of these 138 abstracts originate from the LSF200 NER corpus, which focused on LSFs and did not require any diseases to be mentioned. Only 29 abstracts have no LSF mentions. This too is primarily because of LSF200, in which documents were selected randomly from specific journals. However, a portion of abstracts without entity mentions come from the additional 400 abstracts, not just LSF200. Despite requiring at least five LSF and five disease mentions pre-annotated by the JensenLab Tagger, some abstracts have zero mentions after manual correction of false positives.

### Relation extraction

#### System evaluation

We fine-tuned the RoBERTa-large-PM-M3-Voc-hf model on the development set using a grid search of the parameter combinations in Supplementary Table 5. After each epoch, the evaluation is checked, and only the best performing epoch is reported. The best hyperparameters were Maximum Sequence Length (MSL)=180, Learning Rate (LR)=5e-5, and Maximum number of training epochs=75. This resulted in 75.5% precision, 65.2% recall, and 70.0% F-score on the development set. The best model was used for the final prediction on the held-out test set, yielding 77.3% precision, 61.6% recall, and 68.5% F-score. The performance on individual LSF–disease relation types is shown in Figure 5.

**Figure 5.**
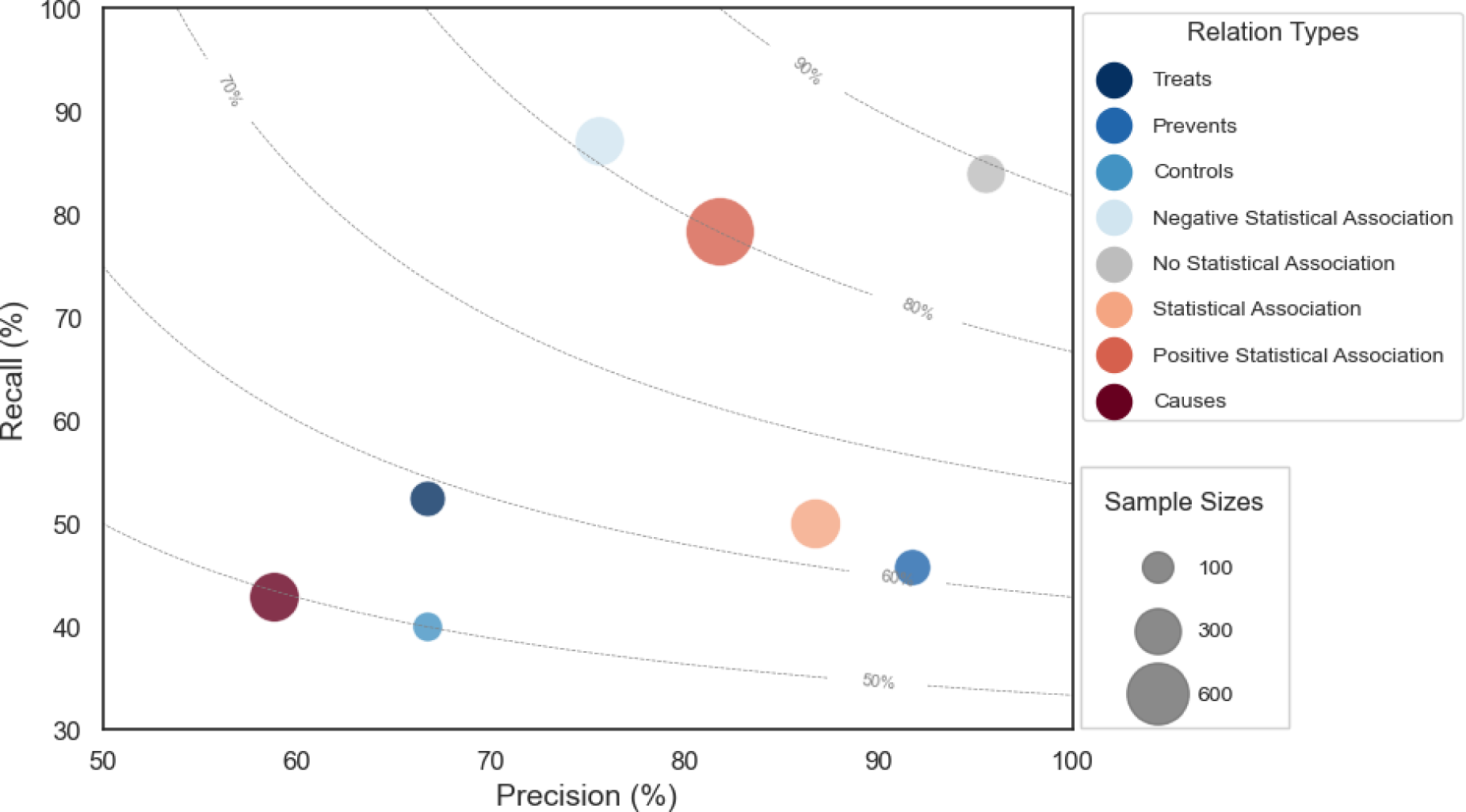
Performance plot for relation extraction across different relation types. Marker size corresponds to the number of examples in the corpus. The dotted lines represent different F-score contours. The size of each circle represents the number of relations in the corpus, and the color corresponds to each of the eight different relation types.

#### Manual error analysis

We manually classified the errors made by the best model on the test set in seven categories (Table 2). The most prevalent category of errors is *Cross-sentence relations*, which refers to any error that crosses sentence boundaries. The model fails to extract 61 of the 77 such relations in the test set, which accounts for more than a quarter of all errors and 40% of the FNs. This problem is not primarily due to the maximum sequence length of the model — which only 8 examples exceed — but is rather due to cross-sentence RE being inherently hard for current models (29,30).

**Table 2.**
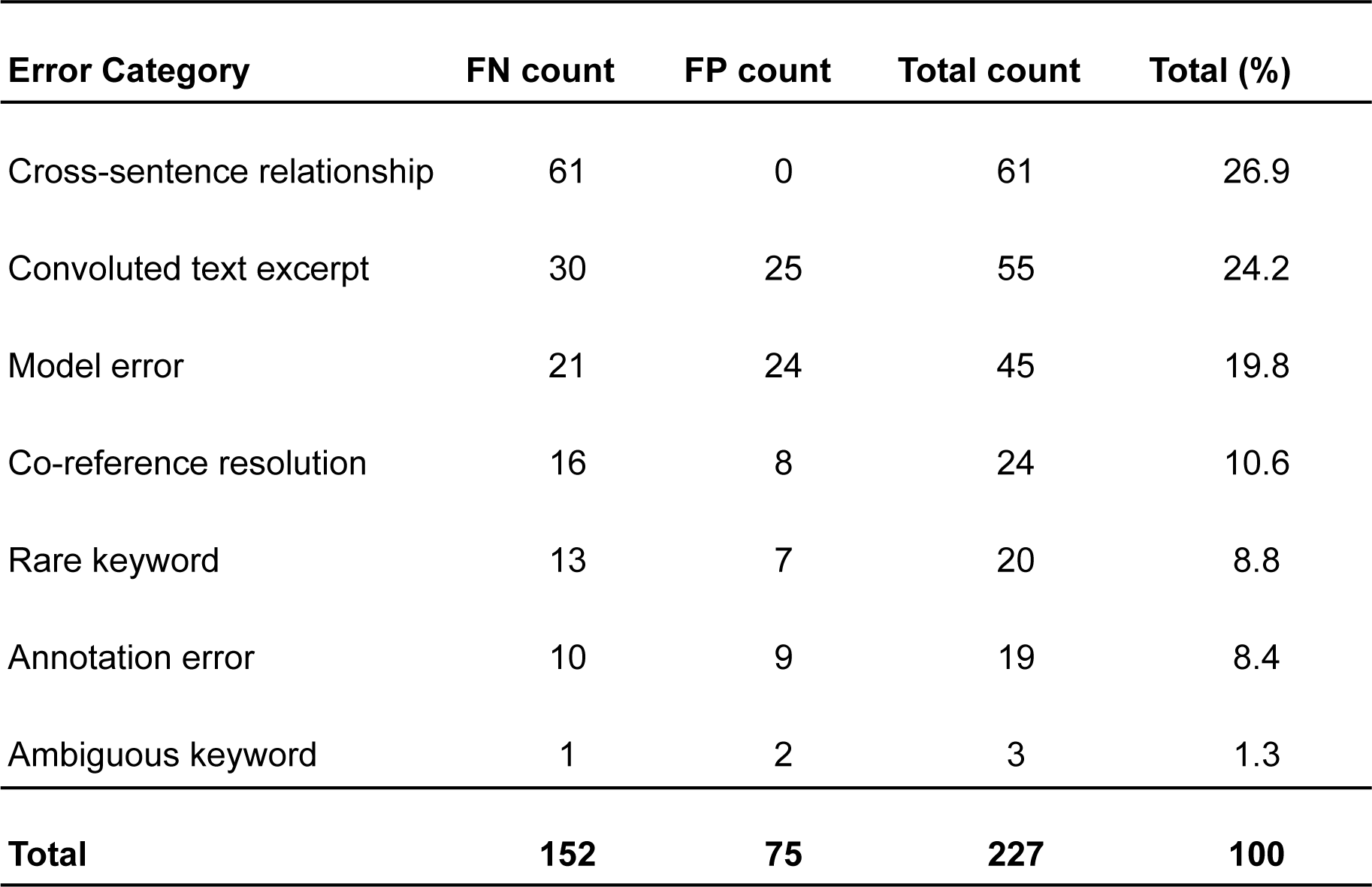
Manual error analysis on the held-out test set.

*Convoluted text excerpt* refers to sentences that are written in a way that makes them challenging to understand also for a human. *Co-reference resolution* errors are closely related to *convoluted text excerpt* errors, but refer specifically to cases where determiners (e.g. *this*, *its*) cause difficulty in identifying which entities are being discussed. *Ambiguous keywords* refers to sentences in which the model likely got confused by words/phrases that can have multiple meanings. For example, in the sentence “*Dietary management is important for those with celiac disease”,* the word *“important*” does not give an unambiguous clue to the relation type. *Rare keyword* errors are caused by words/phrases that have a clear meaning but for which there were insufficient training examples for the model to learn them. *Model errors* comprise wrong predictions that do not belong to any of the categories above. For example, the model could be asserting hypotheses as a fact or simply misinterpreting perfectly comprehensible phrases.

Lastly, we found only 19 *annotation errors* where manual inspection revealed that the model’s predictions are correct and the corpus annotations were wrong. Correcting these, and recalculating the performance metrics increases the F-score by 2.1%.

Another way to understand the errors made by the model is to look whether different relation types get confused with each other, and if so, which ones get confused. The vast majority (82%) of FNs are not caused by label confusion, but are simply completely missed by the model. Almost half of these are cross-sentence relations. Similarly, 63% of the FPs are relations predicted by the model where there should be no relation.

Relation types being confused with each other accounts for 55 out of 227 (24%) of all errors (FP+FN). The relations more commonly confused are *Prevents* being predicted as *Negative Statistical Association*, *Statistical Association* being predicted as *Causes*, and *Causes* being predicted as *Positive Statistical Association*.

Although accurate relation extraction at the most specific level of granularity is ideal, a RE system is already useful if it can identify whether a relation exists or not and whether it is positive or negative. At this level of granularity, only 8 of the 55 remain and the F-score on the test set improves by 4.9% from 68.5% to 73.4%.

#### Validation results on external corpora

To further validate the RE system, we extended our evaluation beyond the held-out test set from the LSD600 corpus to include the only publicly available corpora for lifestyle factors: the Nutrition and Disease (ND) corpus and the FoodDisease corpus. Despite being limited to nutrition and not being perfectly aligned with our corpus in terms of relation types, these corpora provided a valuable benchmark for assessing the generalizability of our system.

To use our RE system to make predictions on these corpora, we mapped the relation types from LSD600 to the relation types in each of the two other corpora (see Methods for details). For the ND corpus, the RE system achieved 92.0% precision, 27.3% recall, and 42.2% F-score. To understand the very low recall, we performed an error analysis. Most of the FNs turned out to be relations that should not be annotated according to our guidelines. For example, the ND dataset included hypothetical statements and relations between diseases and metabolic biomarkers. When excluding the FNs that fall outside the scope of our annotation guidelines, the recall is 57.5% and the F-score is 70.7%.

For the FoodDisease corpus, the RE system achieved a precision of 94.3%, and a recall of 54.8%, resulting in an F-score of 69.3%. Similar to the ND corpus, an error analysis was performed to exclude false negatives due to out-of-scope relations. This led to a 70.7% recall and 80.7% F-score.

The excellent precision achieved on both corpora indicates that the relations identified by our system as LSF–disease relations are perfectly accepted by the standards of these other corpora. These performance results are especially promising considering that we did not train our model on these external datasets but used the model trained on LSD600.

### Conclusions

In this study, we present LSD600, the first of its kind, manually-curated, LSF–disease corpus for relation extraction. We believe LSD600 will be an invaluable resource for the development and benchmarking of RE methods. In contrast to existing corpora, LSD600 encompasses all aspects of LSFs and annotates eight different relation types between these LSFs and diseases. Our annotations are also not limited to sentence boundaries in the text and include 16% cross-sentence relations. Additionally, we present an LSF–disease RE system trained using the LSD600 corpus, which achieved an F-score of 68.5% on the held-out test set. We further validated the model on external corpora, achieving promising performance. This RE system can be a valuable tool for extracting LSF–disease relations and has many downstream applications, given the crucial role of LSFs in disease onset, development, and management.

## Data Availability

The LSF600 corpus and the fine-tuned transformer-based RE system are available under open licenses. The LSF600 corpus, including annotation guidelines, can be accessed on Zenodo at https://zenodo.org/records/12804856. The implementation source code for the RE system is available on GitHub at https://github.com/EsmaeilNourani/LSF_Disease_RE.

## Data Availability

https://zenodo.org/records/12804856

https://github.com/EsmaeilNourani/LSF_Disease_RE

## Supplementary Material

**Table 1:**
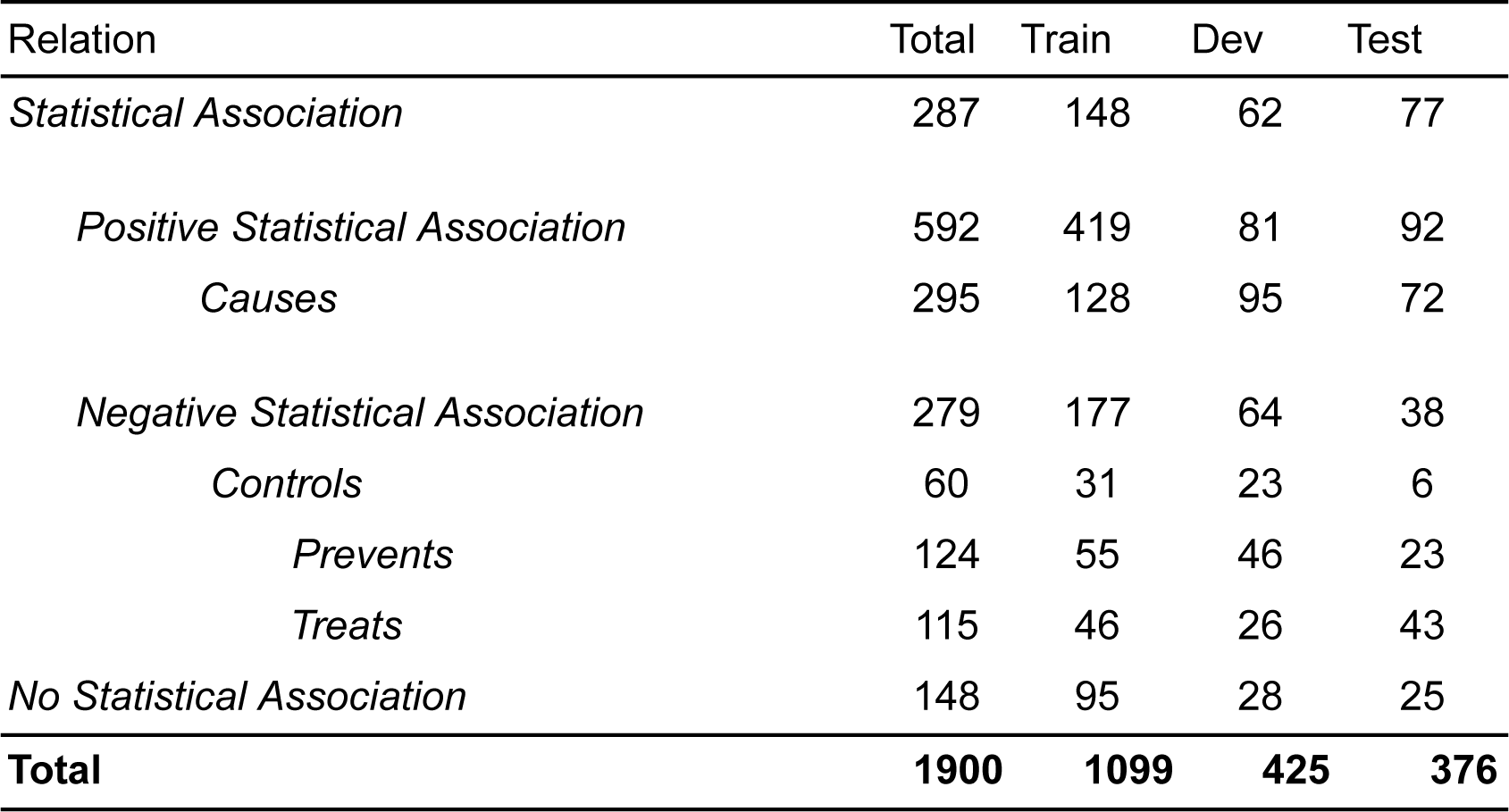
Corpus statistics.

| Relation | Total | Train | Dev | Test |
| --- | --- | --- | --- | --- |
| *Statistical Association* | 287 | 148 | 62 | 77 |
| *Positive Statistical Association* | 592 | 419 | 81 | 92 |
| *Causes* | 295 | 128 | 95 | 72 |
| *Negative Statistical Association* | 279 | 177 | 64 | 38 |
| *Controls* | 60 | 31 | 23 | 6 |
| *Prevents* | 124 | 55 | 46 | 23 |
| *Treats* | 115 | 46 | 26 | 43 |
| *No Statistical Association* | 148 | 95 | 28 | 25 |
| **Total** | **1900** | **1099** | **425** | **376** |

**Table 2.**
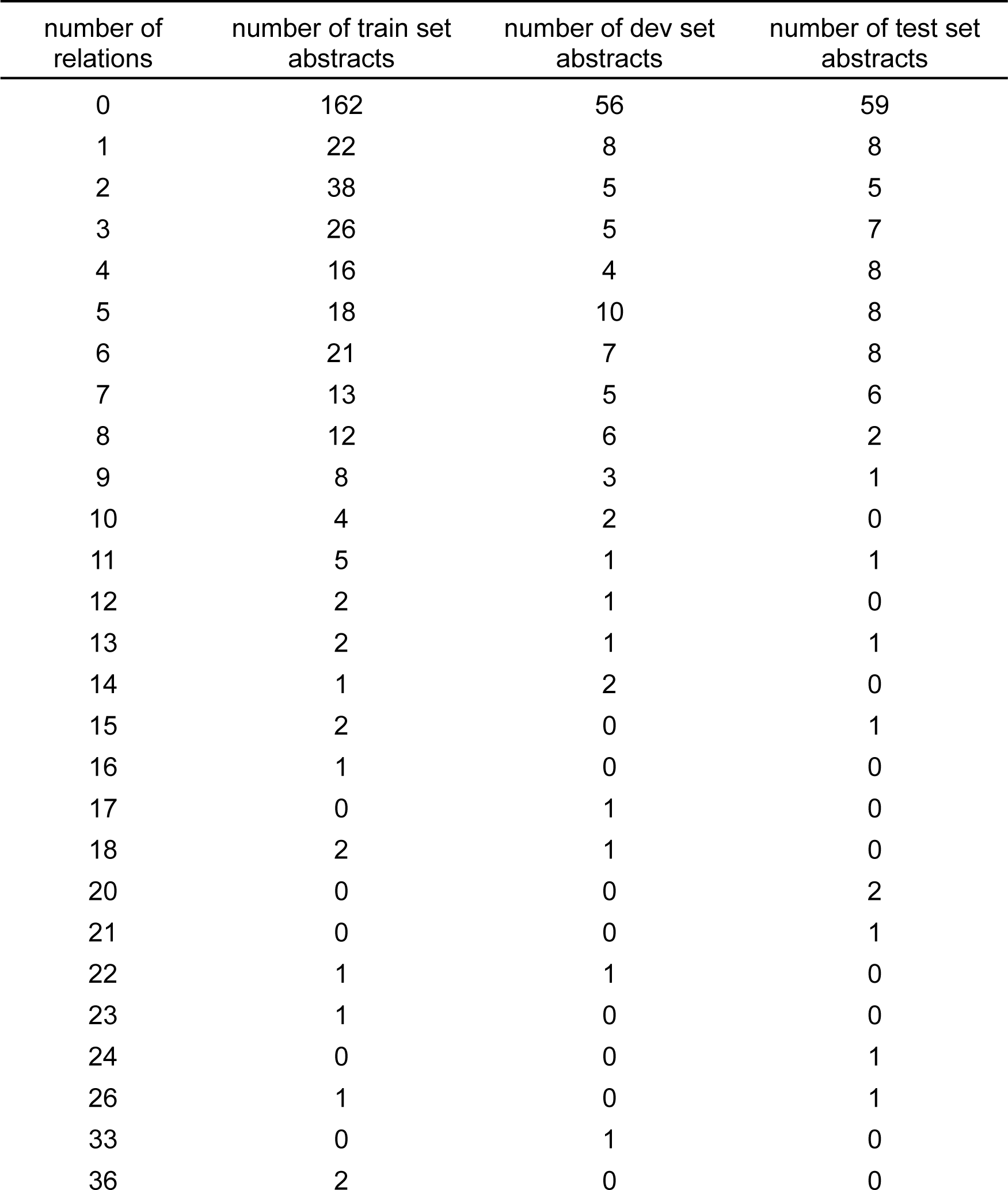
Distribution of relations in train, development and test sets.

**Table 3.** Distribution of Lifestyle Factor mentions in train, development and test sets.

| number of LSF<br>mentions | number of train set<br>abstracts | number of dev set<br>abstracts | number of test set<br>abstracts |
| --- | --- | --- | --- |
| 0 | 17 | 6 | 6 |
| 1 | 13 | 2 | 6 |
| 2 | 11 | 3 | 5 |
| 3 | 19 | 2 | 6 |
| 4 | 8 | 4 | 4 |
| 5 | 17 | 5 | 9 |
| 6 | 10 | 8 | 2 |
| 7 | 23 | 9 | 7 |
| 8 | 21 | 4 | 7 |
| 9 | 25 | 6 | 7 |
| 10 | 19 | 5 | 6 |
| 11 | 14 | 8 | 5 |
| 12 | 23 | 5 | 4 |
| 13 | 18 | 7 | 6 |
| 14 | 11 | 2 | 5 |
| 15 | 19 | 6 | 6 |
| 16 | 14 | 3 | 5 |
| 17 | 12 | 5 | 2 |
| 18 | 12 | 4 | 0 |
| 19 | 7 | 1 | 1 |
| 20 | 14 | 4 | 6 |
| 21 | 5 | 5 | 1 |
| 22 | 3 | 2 | 2 |
| 23 | 4 | 3 | 2 |
| 24 | 5 | 1 | 0 |
| 25 | 2 | 2 | 2 |
| 26 | 3 | 1 | 3 |
| 27 | 3 | 0 | 1 |
| 28 | 0 | 1 | 0 |
| 29 | 2 | 1 | 0 |
| 30 | 2 | 1 | 1 |
| 31 | 0 | 0 | 1 |
| 32 | 0 | 1 | 0 |
| 35 | 1 | 0 | 0 |
| 37 | 1 | 1 | 0 |
| 43 | 1 | 0 | 0 |
| 46 | 1 | 1 | 0 |
| 48 | 0 | 1 | 1 |
| 51 | 0 | 0 | 1 |

**Table 4.**
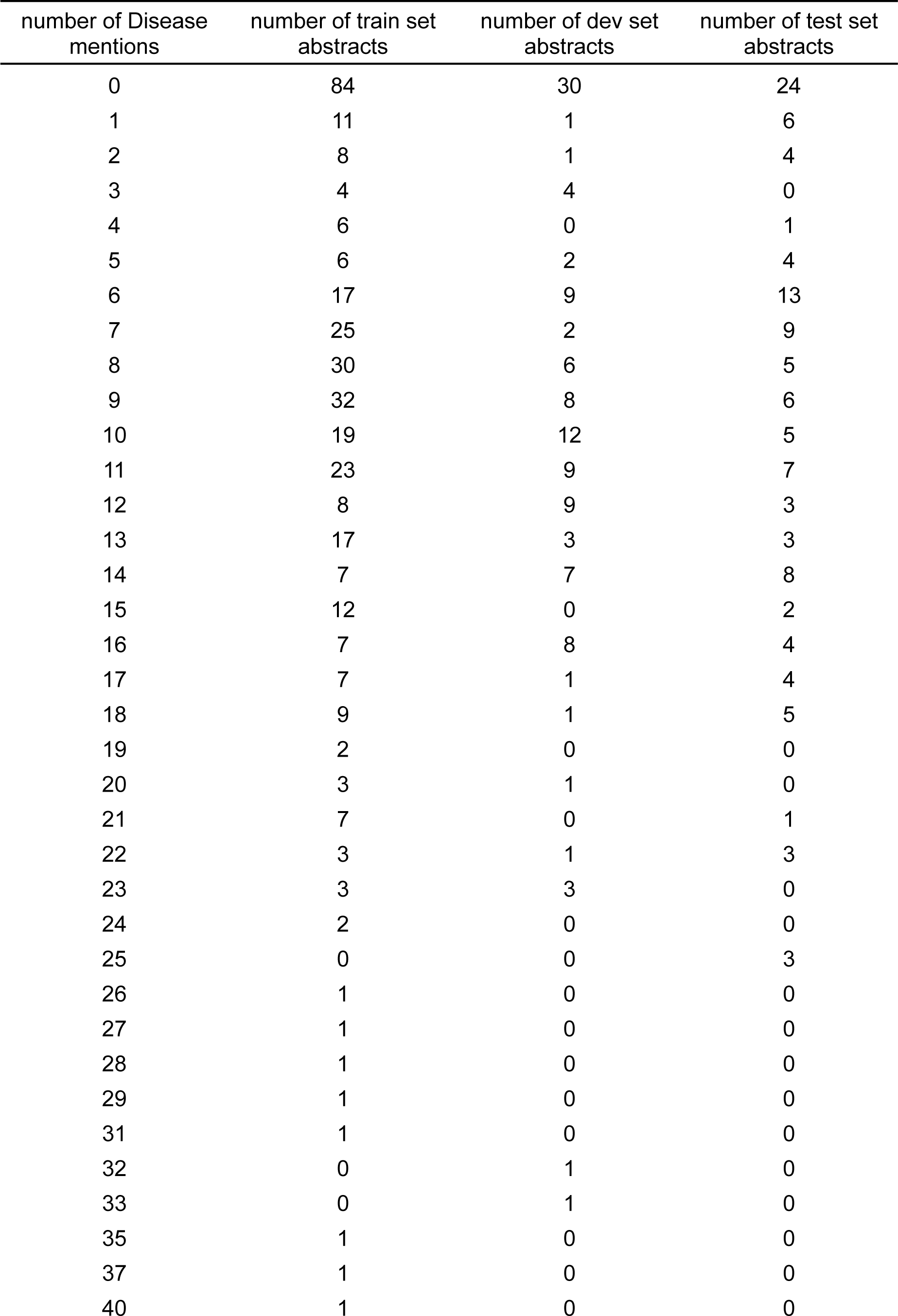
Distribution of Disease mentions in train, development and test sets.

| number of Disease mentions | number of train set abstracts | number of dev set abstracts | number of test set abstracts |
| --- | --- | --- | --- |
| 0 | 84 | 30 | 24 |
| 1 | 11 | 1 | 6 |
| 2 | 8 | 1 | 4 |
| 3 | 4 | 4 | 0 |
| 4 | 6 | 0 | 1 |
| 5 | 6 | 2 | 4 |
| 6 | 17 | 9 | 13 |
| 7 | 25 | 2 | 9 |
| 8 | 30 | 6 | 5 |
| 9 | 32 | 8 | 6 |
| 10 | 19 | 12 | 5 |
| 11 | 23 | 9 | 7 |
| 12 | 8 | 9 | 3 |
| 13 | 17 | 3 | 3 |
| 14 | 7 | 7 | 8 |
| 15 | 12 | 0 | 2 |
| 16 | 7 | 8 | 4 |
| 17 | 7 | 1 | 4 |
| 18 | 9 | 1 | 5 |
| 19 | 2 | 0 | 0 |
| 20 | 3 | 1 | 0 |
| 21 | 7 | 0 | 1 |
| 22 | 3 | 1 | 3 |
| 23 | 3 | 3 | 0 |
| 24 | 2 | 0 | 0 |
| 25 | 0 | 0 | 3 |
| 26 | 1 | 0 | 0 |
| 27 | 1 | 0 | 0 |
| 28 | 1 | 0 | 0 |
| 29 | 1 | 0 | 0 |
| 31 | 1 | 0 | 0 |
| 32 | 0 | 1 | 0 |
| 33 | 0 | 1 | 0 |
| 35 | 1 | 0 | 0 |
| 37 | 1 | 0 | 0 |
| 40 | 1 | 0 | 0 |

**Table 5.** Performance of the grid search on the development set, including hyperparameter values. The best model (highlighted in gray) is used to perform a run on the held-out test set.

| MSL | LR | MaxEpochs | Precision | Recall | F-Score |
| --- | --- | --- | --- | --- | --- |
| 180 | 5e-6 | 75 | **75.45** | **65.15** | **70.0** |
| 180 | 4e-6 | 75 | 70.25 | 66.15 | 68.0 |
| 128 | 5e-6 | 75 | 74.3 | 62.8 | 68.0 |
| 180 | 3e-6 | 75 | 72.15 | 60.45 | 65.8 |
| 180 | 5e-6 | 50 | 71.55 | 60.55 | 65.6 |
| 128 | 3e-6 | 75 | 72.35 | 59.5 | 65.3 |
| 128 | 4e-6 | 75 | 69.95 | 58.2 | 63.5 |
| 180 | 3e-6 | 50 | 64.85 | 60.5 | 62.5 |
| 128 | 2e-6 | 75 | 67.1 | 51.75 | 58.5 |
| 180 | 2e-6 | 75 | 58.9 | 53.9 | 56.3 |
| 128 | 1e-6 | 75 | 61.85 | 50.6 | 55.7 |

## References

1. Byrne, S., Boyle, T., Ahmed, M., et al. (2023) Lifestyle, genetic risk and incidence of cancer: a prospective cohort study of 13 cancer types. International Journal of Epidemiology, 52, 817–826.

2. Olsson, T., Barcellos, L.F. and Alfredsson, L. (2017) Interactions between genetic, lifestyle and environmental risk factors for multiple sclerosis. Nat Rev Neurol, 13, 25–36.

3. Sun, Y., Yuan, S., Chen, X., et al. (2023) The Contribution of Genetic Risk and Lifestyle Factors in the Development of Adult-Onset Inflammatory Bowel Disease: A Prospective Cohort Study. Official journal of the American College of Gastroenterology *| ACG*, 118, 511.

4. Said, M.A., van de Vegte, Y.J., Zafar, M.M., et al. (2019) Contributions of Interactions Between Lifestyle and Genetics on Coronary Artery Disease Risk. Curr Cardiol Rep, 21, 89.

5. Mutie, P.M., Giordano, G.N. and Franks, P.W. (2017) Lifestyle precision medicine: the next generation in type 2 diabetes prevention? BMC Med, 15, 171.

6. Mansour, S., Alkhaaldi, S.M.I., Sammanasunathan, A.F., et al. (2024) Precision Nutrition Unveiled: Gene–Nutrient Interactions, Microbiota Dynamics, and Lifestyle Factors in Obesity Management. Nutrients, 16, 581.

7. Fu, C., Pan, X., Wu, J., et al. (2023) KG4NH: A Comprehensive Knowledge Graph for Question Answering in Dietary Nutrition and Human Health. *IEEE J Biomed Health Inform*, **PP**.

8. Dang, L.D., Phan, U.T.P. and Nguyen, N.T.H. (2023) GENA: A knowledge graph for nutrition and mental health. J Biomed Inform, 145, 104460.

9. Cenikj, G., Strojnik, L., Angelski, R., et al. (2023) From language models to large-scale food and biomedical knowledge graphs. Sci Rep, 13, 7815.

10. Grissa, D., Junge, A., Oprea, T.I., et al. (2022) Diseases 2.0: a weekly updated database of disease–gene associations from text mining and data integration. Database (Oxford), 2022, baac019.

11. Kaszuba-Zwoińska, J., Gremba, J., Gałdzińska-Calik, B., et al. (2015) Electromagnetic field induced biological effects in humans. Przegl Lek, 72, 636–641.

12. Watson, M., Holman, D.M. and Maguire-Eisen, M. (2016) Ultraviolet Radiation Exposure and Its Impact on Skin Cancer Risk. Seminars in Oncology Nursing, 32, 241–254.

13. Miłowska, K., Grabowska, K. and Gabryelak, T. (2014) Applications of electromagnetic radiation in medicine. Postepy Hig Med Dosw, 68, 473–482.

14. Devlin, J., Chang, M.-W., Lee, K., et al. (2018) BERT: Pre-training of Deep Bidirectional Transformers for Language Understanding. BERT: Pre-training of Deep Bidirectional Transformers for Language Understanding (2018) .

15. Liu, Y., Ott, M., Goyal, N., et al. (2019) RoBERTa: A Robustly Optimized BERT Pretraining Approach. RoBERTa: A Robustly Optimized BERT Pretraining Approach (2019).

16. Yang, X., Yu, Z., Guo, Y., et al. (2021) Clinical Relation Extraction Using Transformer-based Models. Clinical Relation Extraction Using Transformer-based Models (2021) .

17. Su, J., Wu, Y., Ting, H.-F., et al. (2021) RENET2: high-performance full-text gene–disease relation extraction with iterative training data expansion. NAR Genomics and Bioinformatics, 3, lqab062.

18. Doughty, E., Kertesz-Farkas, A., Bodenreider, O., et al. (2011) Toward an automatic method for extracting cancer- and other disease-related point mutations from the biomedical literature. Bioinformatics, 27, 408–415.

19. Li, J., Sun, Y., Johnson, R.J., et al. (2016) BioCreative V CDR task corpus: a resource for chemical disease relation extraction. Database (Oxford), 2016, baw068.

20. Nourani, E., Koutrouli, M., Xie, Y., et al. (2024) Lifestyle factors in the biomedical literature: comprehensive resources for named entity recognition. Lifestyle factors in the biomedical literature: comprehensive resources for named entity recognition (2024), 2024.06.13.598816.

21. Kim, J.-D., Ohta, T., Pyysalo, S., et al. (2009) Overview of BioNLP’09 Shared Task on Event Extraction. In Tsujii, J. (ed.), Proceedings of the BioNLP 2009 Workshop Companion Volume for Shared Task, Association for Computational Linguistics, Boulder, Colorado, pp. 1–9.

22. Neves, M. and Ševa, J. (2021) An extensive review of tools for manual annotation of documents. Briefings in Bioinformatics, 22, 146–163.

23. Stenetorp, P., Pyysalo, S., Topić, G., et al. (2012) brat: a Web-based Tool for NLP-Assisted Text Annotation. In Segond, F. (ed.), Proceedings of the Demonstrations at the 13th Conference of the European Chapter of the Association for Computational Linguistics, Association for Computational Linguistics, Avignon, France, pp. 102–107.

24. Mehryary, F., Nastou, K., Ohta, T., et al. (2024) STRING-ing together protein complexes: corpus and methods for extracting physical protein interactions from the biomedical literature. STRING-ing together protein complexes: corpus and methods for extracting physical protein interactions from the biomedical literature (2024), 2023.12.10.570999.

25. Nastou, K., Mehryary, F., Ohta, T., et al. (2024) RegulaTome: a corpus of typed, directed, and signed relations between biomedical entities in the scientific literature. RegulaTome: a corpus of typed, directed, and signed relations between biomedical entities in the scientific literature (2024), 2024.04.30.591824.

26. Björne, J., Heimonen, J., Ginter, F., et al. (2009) Extracting complex biological events with rich graph-based feature sets. Proceedings of the Workshop on BioNLP Shared Task - BioNLP *’*09, Association for Computational Linguistics, Boulder, Colorado, p. 10.

27. Miranda-Escalada, A., Mehryary, F., Luoma, J., et al. (2023) Overview of DrugProt task at BioCreative VII: data and methods for large-scale text mining and knowledge graph generation of heterogenous chemical–protein relations. Database, 2023, baad080.

28. Mehryary, F., Björne, J., Salakoski, T., et al. (2018) Potent pairing: ensemble of long short-term memory networks and support vector machine for chemical-protein relation extraction. Database (Oxford), 2018, bay120.

29. Yao, Y., Ye, D., Li, P., et al. (2019) DocRED: A Large-Scale Document-Level Relation Extraction Dataset. Proceedings of the 57th Annual Meeting of the Association for Computational Linguistics, Association for Computational Linguistics, Florence, Italy, pp. 764–777.

30. Nachtegael, C., De Stefani, J., Cnudde, A., et al. (2024) DUVEL: an active-learning annotated biomedical corpus for the recognition of oligogenic combinations. Database (Oxford), 2024, baae039.

